# Associations between affective/vegetative neuropsychiatric symptoms and brain morphology in aging people with mild cognitive impairment and Alzheimer’s disease

**DOI:** 10.1101/2022.10.05.22280745

**Authors:** Adriana Cannizzaro, Lucas Ronat, Lyna Mariam El Haffaf, Alexandru Hanganu, the ADNI

**Author notes:** Correspondance to : Alexandru Hanganu, PhD. Centre de Recherche de l’Institut Universitaire de Montréal, 4545 Chemin Queen Mary, H3W 1W4 Montréal, Québec, Canada. **Alzheimer’s Disease Neuroimaging Initiative** Data used in preparation of this article were obtained from the Alzheimer’s Disease Neuroimaging Initiative (ADNI) database (adni.loni.usc.edu). As such, the investigators within the ADNI contributed to the design and implementation of ADNI and/or provided data but did not participate in the analyses or writing of this report. A complete listing of ADNI investigators can be found at: https://adni.loni.usc.edu/wp-content/uploads/how_to_apply/ADNI_Acknowledgement_List.pdf.

## Abstract

**Objectives:** Neuropsychiatric symptoms (NPS) are common in mild cognitive impairment (MCI) and even more so in Alzheimer’s disease (AD). The affective/vegetative NPS cluster model (sleep disorders, depression, appetite changes, anxiety, and apathy) has been associated with an increased risk of dementia in patients with MCI and these five NPS have common neuroanatomical associations. Hence, in this study, we examined how brain morphology is influenced by the severity of affective/vegetative NPS across different stages of cognitive performance.

**Participants:** 175 AD, 367 MCI and 223 cognitively normal (CN) participants.

**Setting:** Participants were recruited at multiple centers in North America included in the ADNI project.

**Design:** A GLM was established to test for intergroup differences (CN -MCI, CN-AD, AD-MCI) of the effects of the five NPS on brain structures. A regression model was also performed to show slope directionality of the regions of interest as NPS severity increases.

**Measurements:** 3T MRI data (cortical volumes, areas and thickness) and severity scores of the five NPS.

**Results:** Associations within AD were predominantly stronger compared to MCI. Increased severity of sleep disorders and appetite changes were associated with a decrease in frontal surface areas in AD. Furthermore, increased severity of all NPS (except apathy) were associated with changes in the temporal regions, predominantly with decreased volumes and surface areas.

**Conclusion:** These findings show the implication of fronto-temporal regions with sleep disorders, depression and appetite changes, and contribute to a better understanding of brain morphological differences between CN, MCI and AD with respect to all five NPS.

## Introduction

Neuropsychiatric symptoms (NPS) are described as behavioral and psychological symptoms and include entities such as depression and apathy (Ford, 2014). The severity and frequency of NPS have been established as predictors in terms of mortality risk and nursing home placement (Tun *et al*., 2007). NPS are highly prevalent in Alzheimer’s disease (AD) (Zhao *et al*., 2016), such that most AD patients have multiple NPS as reported by a meta-analysis (Brodaty *et al*., 2015).

Recent studies that grouped NPS into subsyndromes or clusters (Aalten *et al*., 2003; Cheng *et al*., 2012; Kang *et al*., 2010; Liew, 2019; Sayegh and Knight, 2014) reported that common, co-occurring NPS are highly correlated with each other and could help identify underlying pathological mechanisms in a more appropriate way than studying NPS individually (Cheng *et al*., 2012; Liew, 2019). However, there is still no consensus on clusters since many studies have reported different symptom clusters of NPS (Aalten *et al*., 2003; Connors *et al*., 2018; Liew, 2019; Sayegh and Knight, 2014; Travis Seidl and Massman, 2016). A recent cohort study by Liew (2019) has shown that the NPS cluster model that includes (1) sleep disorders, (2) depression, (3) appetite changes, (4) anxiety and (5) apathy, i.e., affective/vegetative symptoms, was significantly associated with the risk of dementia in patients with mild cognitive impairment (MCI). This cluster of the same five NPS was also reported by confirmatory factor analysis models of Cheng *et al*. (2012) as well as Sayegh and Knight (2014).

These five NPS have been previously associated with brain morphology changes. Specifically, the anterior cingulate cortex (ACC) and temporal regions (mainly middle and inferior gyrus) have both been shown to have associations with affective/vegetative symptoms, despite the scant number of studies with respect to sleep and appetite disorders (Apostolova *et al*., 2007; Boublay *et al*., 2016; Chen *et al*., 2021; Guercio *et al*., 2015; Ismail *et al*., 2008; Johansson *et al*., 2020; Tascone *et al*., 2017; Zahodne *et al*., 2013). More specifically, the presence of these symptoms has been mostly associated with cortical thinning, decreased volume and/or atrophy in the anterior cingulate and temporal regions. There are also smaller groupings of these five NPS that have common regions. Indeed, sleep disorders, depression and apathy have all three been associated with the middle frontal gyrus, where a decrease in volume of the latter was associated with depression specifically (Boublay *et al*., 2016; Bruen *et al*., 2008; Hu *et al*., 2015). Depression has also a common neuroanatomical correlate with anxiety, where both have been associated with decreased entorhinal cortex thickness and volume respectively (Mah *et al*., 2015; Zahodne *et al*., 2013). Moreover, depression and apathy have been associated with the caudate nucleus region (Bruen *et al*., 2008; Chen *et al*., 2021). Changes with respect to the orbitofrontal cortex have been associated with appetite disorders and apathy, particularly in terms of hypoperfusion and atrophy in this region respectively (Ismail *et al*., 2008; Stanton *et al*., 2013). Furthermore, anxiety and apathy have also both been associated with atrophy of the insular cortex (Mohamed Nour *et al*., 2021; Stanton *et al*., 2013).

In addition to common neuroanatomical associations, these five NPS have also shared neurocircuits. Indeed, a fronto-limbic circuit has been found to be implicated in depression and its ventral part consisting of inferior temporal and subgenual cingulate regions has been associated with insomnia (sleep disorders) and loss of appetite (Chen *et al*., 2021). In addition, the ACC and its connected structures play a role in appetite and hunger as well as in emotion regulation and modulation of amygdala responses to anxiety (Drevets *et al*., 2008; Tascone and Bottino, 2013). The ACC is also involved in motivation and goal-directed behavior where the result of a dysfunction in this region has been implicated in apathy (Rosen *et al*., 2005).

Taken together, these findings in the literature show that not only have these five NPS been grouped in the same cluster, but that they also share multiple regions and neurocircuits. Thus, the aim of this study is to provide support to these findings to determine associations between the affective/vegetative NPS and brain morphology as well as shared regions between all or some of these symptoms through intergroup comparisons (CN-MCI, CN-AD, AD-MCI).

Given the aforementioned associations between the affective/vegetative symptoms, specifically, the middle frontal and temporal regions for sleep disorders and depression as well as the ACC for appetite changes, anxiety, and apathy, we expect that we will find the same associations within the MCI and AD group. Moreover, we expect these associations to be stronger in the AD group as prevalence and severity of NPS are known to increase with disease progression (Chen *et al*., 2021).

The current study will thus focus on the mentioned symptoms: sleep disorders, depression, appetite changes, anxiety, and apathy, i.e., affective/vegetative symptoms.

## Methods

### Participants

Data of 175 patients with AD, 367 patients with MCI, and 223 cognitively normal participants (CN) from the ADNI database were extracted. Data used in the preparation of this article were obtained from the ADNI database (adni.loni.usc.edu). The ADNI, launched in 2003 and led by Principal Investigator Michael W. Weiner, MD, has for main objective to understand the progression of MCI and early AD by combining imaging, biological and neuropsychological data (Mueller *et al*., 2005; Shaw *et al*., 2007) (http://www.adni-info.org/). Entry criteria for patients with amnestic MCI include a Mini-Mental State Examination score (MMSE) of 24 to 30 and a Memory Box score of at least 0.5, whereas other details on the ADNI cohort can be found online. All patients with AD met National Institute of Neurological and Communication Disorders/Alzheimer’s Disease and Related Disorders Association criteria for probable AD with a MMSE score between 20 and 26, a global Clinical Dementia Rating of 0.5 or 1, a sum-of-boxes Clinical Dementia Rating of 1.0 to 9.0, and, therefore, are only mildly impaired. Exclusion criteria included any serious neurological disease other than possible AD, any history of brain lesions or head trauma, or psychoactive medication use (including antidepressants, neuroleptics, chronic anxiolytics, or sedative hypnotics). Retained participants had completed a neuropsychiatric examination, a comprehensive neuropsychological assessment and a 3 Tesla MRI. Ethics committee approval and individual patient consents were received by the corresponding registration sites according to ADNI rules (http://adni.loni.usc.edu/methods/documents/). All data are available on the ADNI websites upon demand (http://adni.loni.usc.edu/data-samples/access-data/). This study was approved by the Comité d’éthique de la recherche vieillissement-neuroimagerie CER VN 19-20-06.

### Neuropsychiatric assessment

We used the neuropsychiatric inventory to extract the corresponding neuropsychiatric symptoms, specifically: delusions, hallucinations, agitation/aggressiveness, depression, anxiety, euphoria, apathy, disinhibition, irritability, aberrant motor behavior, sleep and nighttime behavior disorders (sleep disorders) as well as appetite and eating changes (appetite changes). The neuropsychiatric inventory is aimed at evaluating the behavioral changes of patients in relation to their state in the last month previous state. We used the hetero questionnaire to assess behavioral changes from the point of view of relatives. For each symptom, a general question is asked about the presence or absence of the symptom in relation to the patient’s previous state. If the symptom is present, other questions are asked to specify its characteristics. Finally, the respondent is asked to estimate the severity (i.e. the impact it has on daily life) on a scale of 0 to 3. In this study, the severity of each NPS was considered.

### Procedure

The MRIs from the ADNI database (https://ida.loni.usc.edu/) were processed on the Cedar cluster of the Digital Research Alliance of Canada (http://www.alliancecan.ca), on the CentOS Linux version 7, with FreeSurfer 7.1.1 (http://surfer.nmr.mgh.harvard.edu), Centos 7.0_x86_64 version, (Fischl *et al*., 1999; Fischl *et al*., 2004) and automatically managed and verified by an in-house pipeline (github.com/alexhanganu/nimb). The results enabled the obtaining of volume, thickness and area of cortical regions of interest based on the Desikan atlas. Cortical thickness was computed as the average of (1) the distance from each white surface vertex to their corresponding closest point on the pial surface (not necessarily at a pial vertex) and (2) the distance from the corresponding pial vertex to the closest point on the white surface (Fischl and Dale, 2000). Cortical surface area was computed based on the triangular face of the surface representation with corresponding vertex coordinates *abc* of the corresponding triangle corner and dividing by two the vector norm of the cross product x of the differences between vertex coordinates: |*(a-c)* x *(b-c)*| */2* (Winkler *et al*., 2012). Cortical volume was based on defining oblique truncated triangular pyramids which were divided into three irregular tetrahedra and their volumes were calculated (Winkler *et al*., 2018). Each voxel in the normalized brain volume was assigned to one of 40 labels, using a probabilistic atlas obtained from a manually labeled training set (Fischl *et al*., 2002). Brain volumes were corrected using the Estimated Total Intracranial volume which is a metric computed from the amount of scaling based on the MNI305 space talairach transformation (Buckner *et al*., 2004). We used the regression-based correctional method, which was shown to provide advantages over the proportion method (Sanfilipo *et al*., 2004).

Python 3.9 with Pandas 1.1.2 library was used to compile data of participants. Participants were excluded from analysis due to: (1) incomplete neuropsychiatric evaluations, (2) absence of a 3T MRI, (3) presence of psychiatric (major depression, schizophrenia, bipolar disorder, substance abuse, post-traumatic stress, obsessive-compulsive disorder) or neurological (stroke, head trauma, brain tumor, anoxia, epilepsy, alcohol dependence and Korsakoff, neurodevelopmental disorder) history, (4) absence of prematurity, (5) presence of diagnostic criteria in favor of another neurodegenerative or neurological etiology in the broad sense (clinical, physiological or genetic criteria for Parkinson’s disease, frontotemporal degeneration, progressive supranuclear paralysis, corticobasal degeneration, Lewy body dementia, amyotrophic lateral sclerosis, multiple sclerosis, multi-system atrophy, vascular dementia).

### Statistical analysis

For the descriptive analyses, the data was analyzed using SPSS version 26 software to compare the three groups in terms of their female-male ratio (Pearson’s chi-squared test) and in terms of age, years of education, MMSE scores and prevalence of the affective/vegetative NPS means (univariate analysis of variance and post hoc tests with Bonferroni correction). In addition, the pipeline developed by the research team made it possible to carry out analyses based on the general linear model (GLM) in order to define the neuropsychiatric variables having effects on brain structures, and to obtain the cerebral images from the Monte-Carlo correction. The severity of NPI subscale and the NPI total score were included as fixed factors, while MRI variables (cortical volume, thickness and areas) were regarded as dependent variables. For all the tests, the alpha level was set at 0.05 and the Max-value indicates the difference in slope of volume/surface area/thickness relative to NPS severity between two groups (CN-MCI, CN-AD and AD-MCI). Given that the volume of the cortex is ultimately determined by area and thickness, which are correspondingly influenced by different factors (Panizzon *et al*., 2009; Winkler *et al*., 2010), we analyzed our results in light of volumetric changes as a main change, while thickness and area were considered for secondary explanation.

## Results

Demographic data is shown in Table 1. The MCI (n=367) group had a higher percentage of men than the AD (n=175) and the CN (n=223) groups. Groups did not differ significantly in age. The AD group did have a lower education than the MCI and the CN groups. As expected, MMSE scores were lower for AD than MCI and for MCI than CN. The AD group had a higher prevalence of sleep disorders, depression, appetite changes, anxiety and apathy compared to the MCI group and the latter had a higher prevalence of these NPS than the CN group. For NPS severity information per group, see Table 1/ appendix A1 (published as supplementary material online attached to the electronic version of this paper at https://www.cambridge.org/core/journals/international-psychogeriatrics). The AD group had proportionally higher severity for all five NPS than the MCI group and the same for the MCI group compared to the CN group.

**Table 1.** Sociodemographic data.

| **Characteristics** | **CN**<br>(n= 223) | **MCI**<br>(n=367) | **AD**<br>(n=175) | **P value** |
| --- | --- | --- | --- | --- |
| Female: Male ratio (%) | 48.4: 51.6 | 36.0: 64.0 | 48.6: 51.4 | 0.002 |
| Age (years) | 75.8± 5.1 | 74.8 ± 7.4 | 75.5 ± 7.5 | 0.2 |
| Education (years) | 16.0 ± 2.9 | 15.7± 3.0 | 14.7± 3.1 | <.001*§ |
| MMSE score | 29.1 ± 1.0 | 27.1± 1.8 | 23.5± 2.0 | <.001*†§ |
| Sleep disorders | 20 (9.0%) | 41 (11%) | 42 (24%) | <.001*§ |
| Depression | 12 (5.4%) | 68 (19%) | 57 (33%) | <.001*†§ |
| Appetite changes | 1 (0.4%) | 41 (11%) | 28 (16%) | <.0.01†§ |
| Anxiety | 8 (3.6%) | 63 (17%) | 56 (32%) | <.001*†§ |
| Apathy | 3 (1.3%) | 48 (13%) | 59 (34%) | <.001*†§ |

*Results are expressed as mean ± standard deviation and as NPS prevalence for the five symptoms. n observed number of cases.*
\* Posthoc difference (P<0.05) between AD and MCI.
† Posthoc difference (P<0.05) between CN and MCI.
§ Posthoc difference (P<0.05) between CN and AD.

### GLM analysis results

#### Intergroup CN-MCI

CN-MCI intergroup results are presented in Table 2 and Figure 1. Sleep disorders severity was shown to have associations with the temporal regions. Specifically, CN had a significantly stronger association between sleep disorders severity and the volume of the left temporal banks superior and temporal pole than MCI. Additional associations were found in the right fusiform region with changes only with respect to thickness.

**Table 2.**
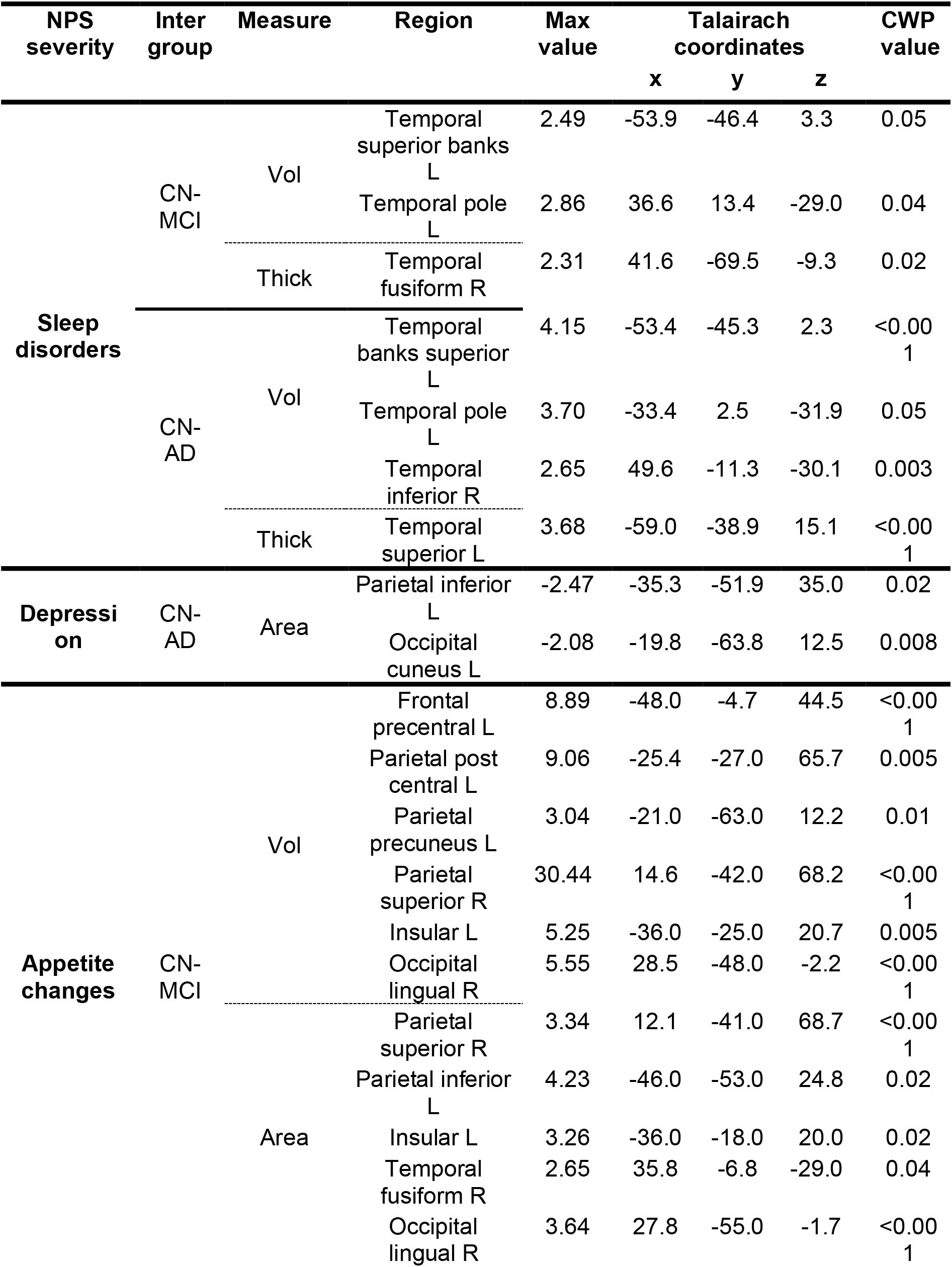
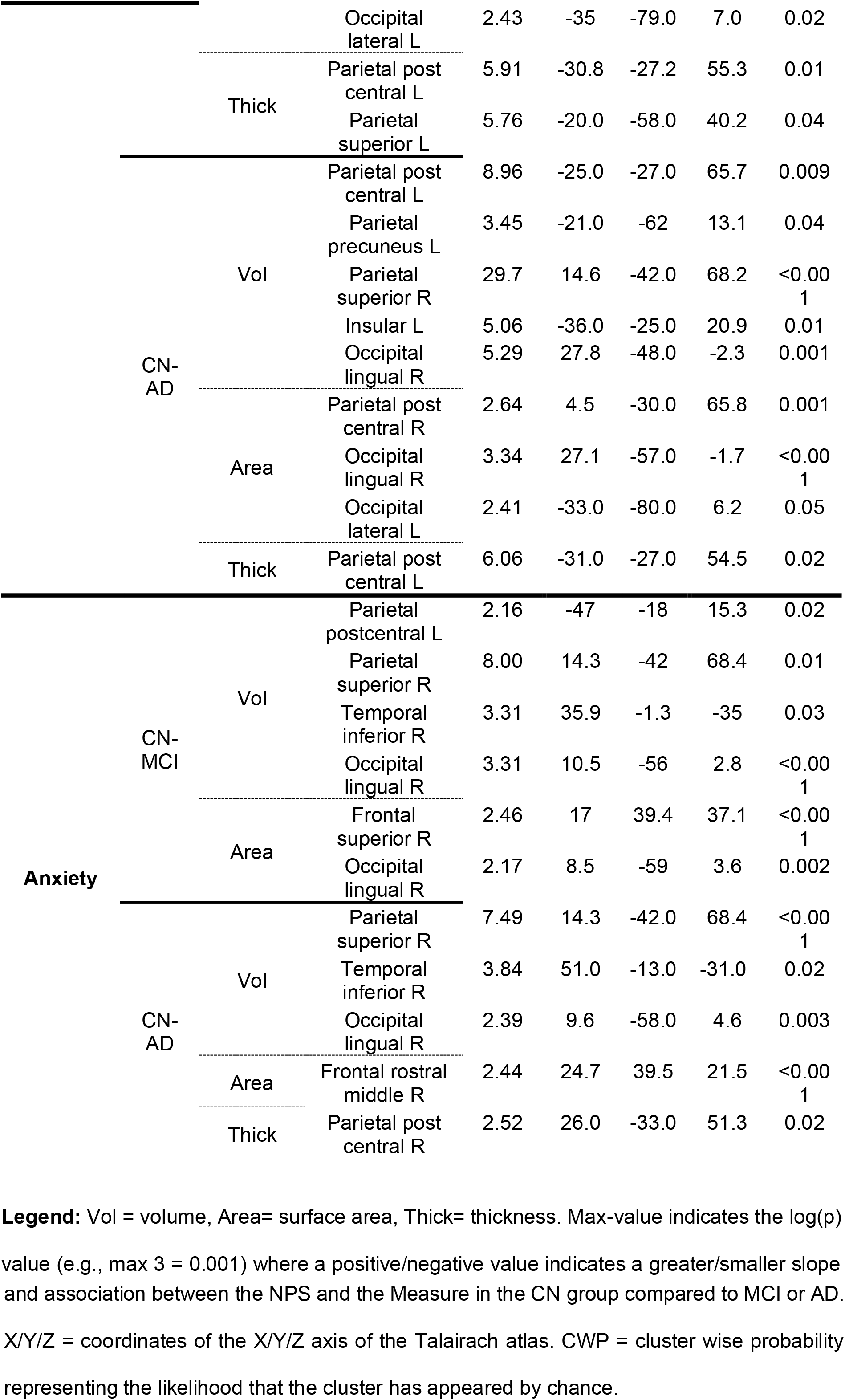
GLM: CN-MCI and CN-AD intergroup results.

**Figure 1.**
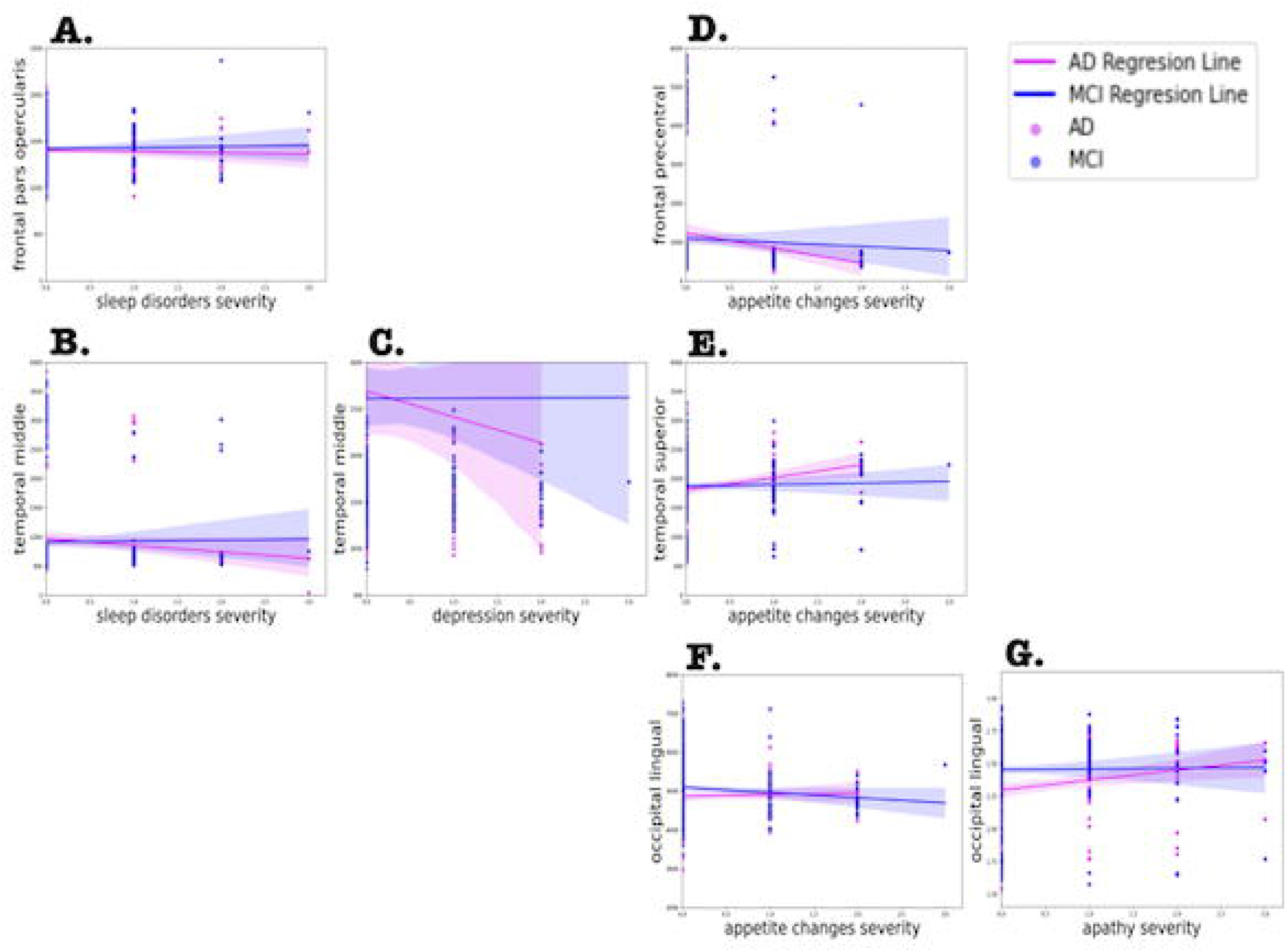
Significant brain structures results of CN-MCI intergroup analysis results of appetite changes severity and the left parietal post central volume (image A, in blue) as well as of sleep disorders severity and the right temporal fusiform thickness (image B). CN-AD intergroup results of anxiety severity and the right parietal superior volume (Image C, in red) and of sleep disorders severity and the right temporal inferior volume (image D, in green). AD-MCI intergroup results of depression severity and the right temporal middle volume (image E, in indigo) as well as of apathy severity and the right occipital lingual thickness (Image F, in purple).

There were no statistically significant results for depression severity for CN-MCI intergroup comparison.

The severity of appetite changes was shown to have associations with the frontal (left precentral), parietal (left post central, left precuneus and right superior), left insular and right lingual regions. Specifically, CN had a stronger volume association with these regions, where most of them had also changes in the surface area and thickness than MCI. Additional associations were found in the left parietal inferior, right temporal fusiform and left occipital lateral regions with changes in the surface area only as well as in the left parietal superior with respect to thickness.

Anxiety severity was shown to have a stronger association with the parietal (left post central, right superior,), right temporal inferior, and right occipital lingual regions within the CN group compared to the MCI group. Similarly, other associations were found in the right frontal superior and right occipital lingual with changes with respect to surface area.

There were no statistically significant results for apathy severity for CN-MCI intergroup comparison.

#### Intergroup CN-AD

Intergroup CN-AD results are presented Table 2 and Figure 1. Sleep disorders severity was shown to have associations with the temporal regions. Specifically, CN had a significantly stronger association between sleep disorders severity and left banks superior, right inferior and left pole with respect to volume than AD. An additional association was found with the left temporal superior thickness.

Depression severity, in contrast, was shown to have associations with the parietal and occipital regions, in the left hemisphere. Specifically, AD had a significantly stronger association between depression severity and the surface areas of the parietal inferior and occipital cuneus regions compared to CN.

The severity of appetite changes was shown to have a stronger association with parietal, insular and occipital regions in the CN group compared to the AD group. In particular, CN had a significantly stronger association between appetite changes severity and the volumes of the parietal regions (left post central, left precuneus, right superior), left insular and right lingual regions, driven mainly by changes in the surface area and thickness than AD.

Anxiety severity was shown to have associations in all four lobes, all in the right hemisphere. CN had a significantly stronger association between anxiety severity and the volumes of the parietal superior, temporal inferior and occipital lingual regions in comparison to MCI. Additional associations were found with the frontal rostral middle and parietal post central surface area and thickness respectively.

There were no statistically significant results for apathy severity for CN-AD intergroup comparison.

#### Intergroup AD-MCI

Intergroup AD-MCI results are shown in Table 3 and Figure 1. Sleep disorders severity was shown to have associations with brain morphology in frontal and temporal regions (Table 3). More specifically, MCI had a significantly stronger association between sleep disorders severity and the volume of left inferior temporal as well as the right par opercularis and left middle temporal surface areas than AD.

**Table 3.**
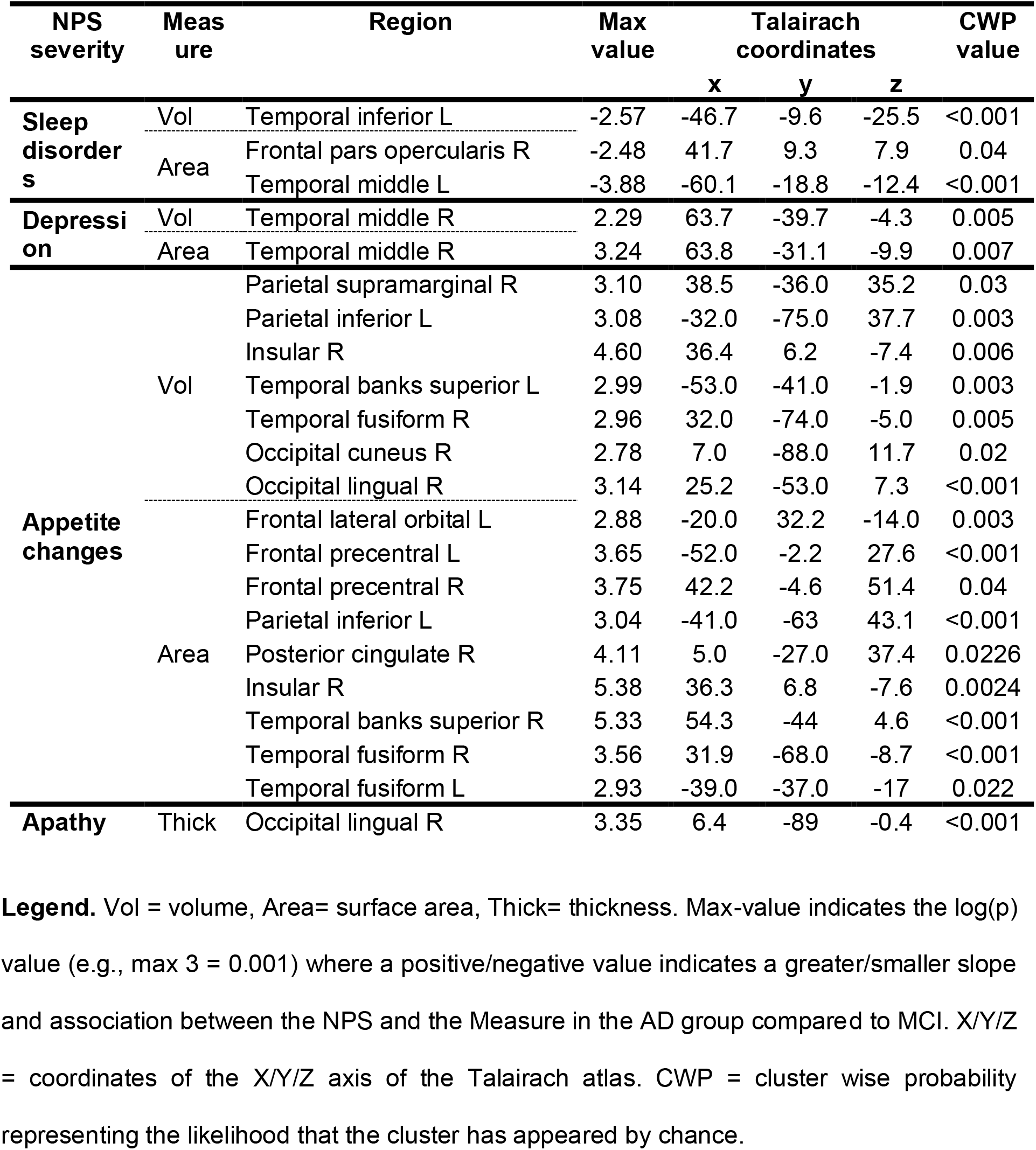
GLM: AD-MCI intergroup results.

Depression severity was shown to have associations with the right temporal lobe. Particularly, AD had a significantly stronger association between depression severity and the volume of the temporal middle region driven by changes in the surface area compared to MCI.

Appetite changes severity was shown to have associations with all lobes. Specifically, AD had a significantly stronger association between appetite changes severity and the volume of the parietal (right supramarginal and left inferior), right insular, temporal (left temporal banks superior and right fusiform gyrus), occipital (right cuneus and lingual) regions than MCI. The parietal inferior, insular and temporal fusiform were driven mainly by changes in the surface area. Additional associations were found with the left frontal lateral orbital, left and right precentral, right posterior cingulate, right temporal banks superior with changes only to surface areas.

There were no statistically significant results for anxiety severity for AD-MCI intergroup comparison.

Apathy severity was shown to have an association only with the occipital region. Particularly, AD had a significantly stronger association between apathy severity and the right occipital lingual thickness than MCI.

AD-MCI regressions images are shown in Figure 2, only the intergroup AD-MCI was retained since the prevalence of higher NPS severity (scores of 2 or 3) among the CN group was too low, or even zero. More particularly, they show that as sleep disorders severity increases, there is a more of a decrease in the right opercularis and left temporal surface area and volume in the AD group than the MCI group. This trend can also be seen with the temporal middle volume and depression severity. By contrast, as appetite changes severity increases, temporal and insular regions increase in the AD group. As for frontal, parietal and occipital regions with appetite changes severity, the results are varied. Lastly, similarly to the trend seen with the temporal regions and appetite changes, in the AD group, as apathy severity increases the right occipital lingual increases too, while this ROI in the MCI group does not change as much.

**Figure 2.**
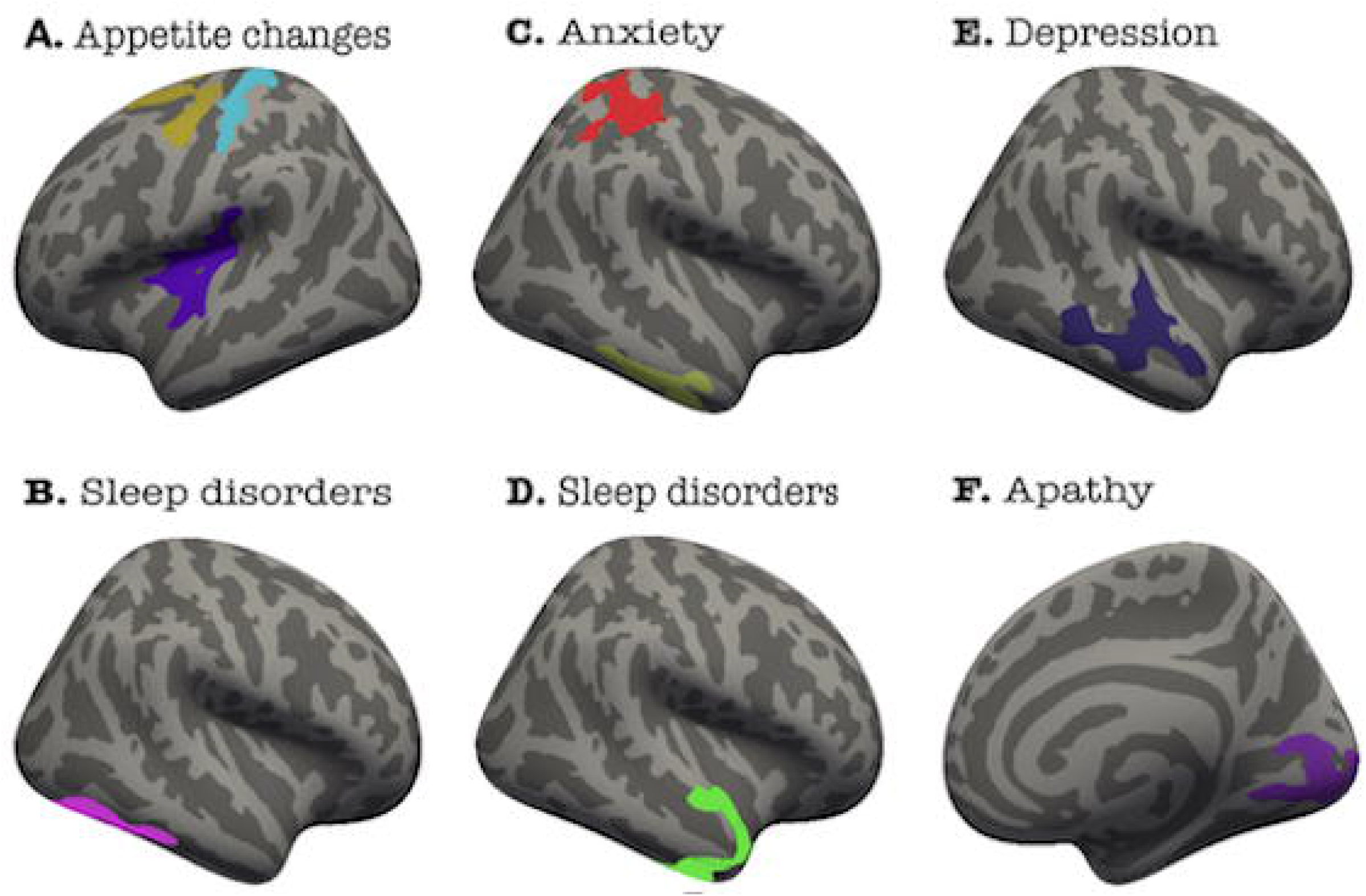
AD-MCI regression results of sleep disorders severity and right frontal par opercularis (image A) and left temporal middle region surface areas (image B). Depression severity and right temporal middle region volume regression result is shown in image C. Appetite changes severity and right frontal precentral surface area (image D), right temporal superior surface area (image E) and right occipital lingual volume (image F). Lastly, apathy and right occipital lingual thickness regression is shown in image G.

## Discussion

Studies have shown that sleep disorders, depression, appetite changes, anxiety and apathy have brain structures and neurocircuits in common. Thus, the aim of this study was to examine associations between severity of these NPS and brain morphology and investigate shared commonalities amongst all or some of them across intergroup comparisons (CN-MCI, CN-AD and AD-MCI). As initially hypothesized, we found that (1) all the affective/vegetative symptoms (except apathy) were significantly associated with temporal regions, (2) associations were predominately stronger in the AD group compared to MCI (3) increased severity of sleep disorders and depression were associated with a decrease in temporal volume and surface areas, (4) sleep disorders, depression and appetite changes were all significantly associated with fronto-temporal regions.

### Sleep disorders

CN-MCI intergroup comparisons revealed that sleep disorders had significant associations with the volume of temporal regions (superior, pole and fusiform). Despite the limited studies on this specific topic, previous studies have also found similar associations with this symptom. Sanchez-Espinosa *et al*. (2017) demonstrated that homocysteine (MCI/AD marker) related temporal volume loss in MCI was dependent on the relationship between sleep problems and reduced antioxidant capacity. In fact, higher levels of homocysteine enhance the neurodegeneration (Obeid and Herrmann, 2006) process and Sanchez-Espinosa *et al*. (2017) stated that poor sleep quality could possibly be an important factor in exacerbating these effects such as temporal atrophy in patients with MCI. Moreover, a recent review (Naismith and Mowszowski, 2018) highlighted studies that have found alterations in networks such as the temporoparietal in patients with MCI and sleep disorders, and these networks are also involved in AD.

CN-AD intergroup comparisons showed the same regions to be significantly associated with sleep disorders in addition to the right temporal inferior.

AD-MCI intergroup comparisons also revealed associations with the temporal regions (inferior, middle) but also with the pars opercularis area. Furthermore, the regression (Figure 2, image B) shows that as sleep disorders severity increases, the left surface area of the middle temporal region decreases. This finding can be supported by Tascone *et al*. (2017) who demonstrated that the presence of sleep disorders in AD correlated with a decreased volume involving the middle temporal pole as well as the middle and inferior temporal regions.

In all, sleep disorders have been associated with the temporal region amongst all stages of cognitive performance (CN, MCI, AD).

### Depression

There were no significant results for depression in the CN-MCI intergroup comparison.

As for the CN-AD, we found associations between depression and the surface areas of the parietal inferior and occipital cuneus in the left hemisphere. Specifically for the parietal inferior, this finding can be supported by a systematic review by Chen *et al*. (2021) that outlined the brain lesions patterns and the frontal-limbic circuit of depression in AD. The parietal lobe, while not the most frequent, is among the regions affected by depression lesions and was found to have cortical thinning in AD patients with depression compared to those without (Chen *et al*., 2021; Lebedeva *et al*., 2014). Additionally, the parietal inferior has anatomical connections to the dorsolateral prefrontal cortex (DLPFC) and both regions are known to be implicated in dorsal compartment of the frontal-limbic circuit (Chen *et al*., 2021; Mayberg *et al*., 1999). The DLPFC has been shown to be involved in the regulation of negative emotion and Koenigs and Grafman (2009) infer a dysfunction or lesions to DLPFC could cause a defect in regulating negative affect and thus, exacerbating depressive symptoms.

AD-MCI intergroup comparison results revealed an association between depression and the right temporal middle region (volume and surface area) and the regression (Figure 2, image C) shows that as depression severity increases, there is a decrease in the temporal middle region for the AD group. Similar results have also been observed in previous studies, where Siafarikas *et al*. (2021) found a negative association between depression and the superior temporal gyrus in patients with AD and Lebedeva *et al*. (2014) demonstrated depression-associated thinning of the superior and inferior temporal also in patients with AD. In all, depression has been associated with the parietal lobe in CN-AD and the temporal region in AD-MCI.

### Appetite changes

Our CN-MCI results show associations between appetite changes and the frontal precentral, the parietal cortex (post central, precuneus, superior, inferior), the insular, the temporal fusiform and occipital regions (lingual and lateral). The same regions were also implicated in the CN-AD intergroup, except the frontal and temporal regions. Appetite changes are very common in AD with one study in 2015 (Kai *et al*., 2015) reporting 81% of patients with AD have eating and swallowing disturbances. Regarding our findings with the parietal regions, this can be supported by a functional imaging study (Uher *et al*., 2004) on a non-demented population that reported a lower activation in the inferior parietal lobule in patients with eating disorders compared to healthy controls.

Moreover, in addition to the regions found in CN-MCI and CN-AD comparisons, additional frontal regions (lateral orbital, precentral, posterior cingulate) were implicated in AD-MCI. Also, regression of the frontal precentral (Figure 2, image D) shows a decrease in this ROI as appetite changes severity increase. The role of frontal and temporal regions in appetite changes in AD can be supported by a functional imaging study by Ismail *et al*. (2008) that revealed hypoperfusion to the left orbitofrontal and relative sparing in the left middle mesial temporal cortices were significant predictors of appetite loss in patients with AD. When it comes to eating, there are two types of mechanisms related to its motivation. One is the intrinsic factor of appetite, which is the homeostatic processes that induce hunger after a period of not eating, and the other is the extrinsic factor, which is stimuli associated with food that can induce a feeling of hunger and desire even when we are not hungry (Hinton *et al*., 2004). In this view, regarding our findings of the frontal orbital, cingulate posterior and the insular cortices, a critical review by Tascone and Bottino (2013) highlighted the role of ACC and its connected structures (orbitofrontal and insular cortices) in the intrinsic factors of appetite and are involved in circuits that are activated in response to starvation. Furthermore, the orbitofrontal cortex is also a neural substrate of the extrinsic factors where its role is involved in the promotion of appetite and production of goal-directed behaviors in eating (Tascone and Bottino, 2013). Thus, atrophy in these regions could contribute to altered appetite signals such as decrease appetite and weight loss in AD (Smith and Greenwood, 2008). Lastly, the regression of the occipital lingual gyrus (Figure 2, image F) shows an increase in this ROI as appetite changes severity increase. A similar finding was reported by Uher *et al*. (2004) where patients with eating disorders had a greater activation of the occipital lingual gyrus compared to controls. In all, appetite changes have been associated with frontal, parietal and occipital regions across all levels of cognitive performance.

### Anxiety

Our CN-MCI intergroup results revealed significant associations between anxiety and the frontal superior, parietal (post central and superior), temporal inferior and occipital lingual regions. CN-AD results revealed almost identical results, only that the frontal rostral middle region was significant instead of the frontal superior. These results concur with some studies that found that higher anxiety scores correlated with the reduction of volume in parietal regions (precuneus and inferior lobule) and that lesions of anxiety include the frontal lobe, the inferior parietal lobule and superior temporal regions (Chen *et al*., 2021; Tagai *et al*., 2014). More specifically, our findings regarding the associations between anxiety severity and the frontal lobe (superior and rostral middle regions) are partly supported by the amygdala circuit of anxiety in AD. In fact, the amygdala is heavily implicated in anxiety, is known for its role in the interpretation of fear and has neuronal interactions with other regions such as the orbitofrontal cortex which help to initiate coping behaviors (Chen *et al*., 2021). Lastly, Tascone and Bottino (2013) outlined that regions among the prefrontal cortex and temporal lobe which are involved in emotional processing and emotional behaviors have shown abnormalities in adults with anxiety. As for AD-MCI comparison, there were no significant results for anxiety. In all, anxiety has been associated with frontal, parietal post central, parietal superior, temporal inferior and occipital lingual regions across all levels of cognitive performance.

### Apathy

The only significant association for apathy was in the AD-MCI intergroup comparison, and it revealed an association between this symptom and the occipital lingual region. While this finding can be supported by two studies reporting that apathetic patients with AD had greater atrophy and decreased perfusion of the lingual gyrus compared to those without apathy (Benoit *et al*., 2002; Tunnard *et al*., 2011), a review of neuroimaging findings of apathy in AD rather supports the notion of the involvement of frontal-subcortical networks of apathy in AD (Theleritis *et al*., 2014). This network implies an interconnected group of regions structures that include the ACC, the prefrontal cortex, and the basal ganglia and are strongly implicated in the generation of motivation behavior (Le Heron *et al*., 2018; Nobis and Husain, 2018). Findings thus support the association between a disruption of regions in this network and the clinical manifestations of apathy (Le Heron *et al*., 2018). Furthermore, the regression for the occipital lingual (Figure 2, image G) shows an increase in this ROI as apathy severity increase. This is a surprising finding, but a possible explanation to this can be due to the inflammatory process in AD. This neuroinflammation occurs by the activated microglia, astrocytes, stressed neurons and AB plaques which produce proinflammatory cytokines (Rubio-Perez and Morillas-Ruiz, 2012). Lastly, Holmgren *et al*. (2014) found that a trend towards correlation between apathy and higher levels of sIL-1 RII in cerebral spinal fluid, in which they stated that patients with apathy may have higher levels of neuroinflammation.

### Commonalities between NPS of interest

Our results demonstrate that sleep disorders and appetite changes both have associations with the temporal fusiform region in the CN-MCI intergroup comparison. For CN-AD comparisons, sleep disorders and anxiety both share an association with the temporal inferior region. Lastly, for AD-MCI, sleep disorders and depression have the temporal middle region in common and in both respective regressions, as each of these NPS increase in severity, the temporal middle region decreases in the AD group.

In CN-MCI, appetite changes and anxiety have the parietal post central, parietal superior and occipital lingual regions in common. Appetite changes and anxiety also share the three same associations with the regions in CN-AD. Appetite changes and apathy have the occipital lingual region in common and as the severity of these symptoms increase respectively, the region increases in the AD group shown in the AD-MCI regressions.

### Strengths and limitations

This study has several limitations. First, the NPI-Q that was used to evaluate NPS, assesses them only over the previous month and does not take into consideration any NPS occurrence out of that time frame. Second, most patients with AD in this study were in the earlier stage of the disease, thus this can affect results as they may not be representative of the entire AD population. Third, the MRI acquisitions were done in different centers which could possibly create inter-scanner and software variabilities (Lee *et al*., 2012). Finally, no causal inference can be made between NPS and significant brain regions as this is a cross-sectional study. Thus, further longitudinal studies are needed to investigate the relationship between NPS and brain regions of interest. Despite these limitations, this study has also many strengths. One of the strengths is the relatively large sample size which increases the robustness of our findings. In addition, this is one of the first studies to perform intergroup analysis across three groups of cognitive decline, including cognitively normal participants regarding NPS and brain morphology. Specifically, compared to other studies on NPS and their brain correlates, this study demonstrates brain morphological differences between all three cognitive groups (CN-MCI, CN-AD, AD-MCI) regarding NPS severity.

## Conclusion

To summarize, as hypothesized we identified (1) regions that were significantly associated with sleep disorders, depression, appetite changes, anxiety and apathy at different intergroup comparisons (CN-MCI, CN-AD and AD-MCI), and (2) regions commonalities between these NPS. This study also shed light on changes within regions of interest for MCI and AD groups as NPS increases in severity. In all, these findings contribute to a better understanding of brain morphological differences between CN, MCI and AD groups with respect to all five NPS.

## Data Availability

All data are available on the ADNI website upon demand (http://adni.loni.usc.edu/data-samples/access-data/).

## Abbreviations

ADNI: Alzheimer’s Disease Neuroimaging Initiative
MCI: Mild Cognitive Impairment
CN: Cognitively Normal
NPI: Neuropsychiatric Inventory
NPS: Neuropsychiatric Symptoms
GLM: General Linear Model

## Use of ADNI data

*Data collection and sharing for this project was funded by the Alzheimer’s Disease Neuroimaging Initiative (ADNI) (National Institutes of Health Grant U01 AG024904) and DOD ADNI (Department of Defense award number W81XWH-12-2-0012). ADNI is funded by the National Institute on Aging, the National Institute of Biomedical Imaging and Bioengineering, and through generous contributions from the following: AbbVie, Alzheimer’s Association; Alzheimer’s Drug Discovery Foundation; Araclon Biotech; BioClinica, Inc*.; *Biogen; Bristol-Myers Squibb Company; CereSpir, Inc*.; *Cogstate; Eisai Inc*.; *Elan Pharmaceuticals, Inc*.; *Eli Lilly and Company; EuroImmun; F. Hoffmann-La Roche Ltd and its affiliated company Genentech, Inc*.; *Fujirebio; GE Healthcare; IXICO Ltd*.; *Janssen Alzheimer Immunotherapy Research & Development, LLC*.; *Johnson & Johnson Pharmaceutical Research & Development LLC*.; *Lumosity; Lundbeck; Merck & Co*., *Inc*.; *Meso Scale Diagnostics, LLC*.; *NeuroRx Research; Neurotrack Technologies; Novartis Pharmaceuticals Corporation; Pfizer Inc*.; *Piramal Imaging; Servier; Takeda Pharmaceutical Company; and Transition Therapeutics. The Canadian Institutes of Health Research is providing funds to support ADNI clinical sites in Canada. Private sector contributions are facilitated by the Foundation for the National Institutes of Health (www.fnih.org). The grantee organization is the Northern California Institute for Research and Education, and the study is coordinated by the Alzheimer’s Therapeutic Research Institute at the University of Southern California. ADNI data are disseminated by the Laboratory for NeuroImaging at the University of Southern California*.

---

**Table 1/Appendix A1:** NPS severity per group.

