## Supplementary Table 1 for "Associations between affective/vegetative neuropsychiatric symptoms and brain morphology in aging people with mild cognitive impairment and Alzheimer’s disease"

**Appendix 1**

**Table 1**. NPS severity per group

| **NPS severity** | **CN**  (n=223) | **MCI**  (n=367) | **AD**  **(**n=175) |
| --- | --- | --- | --- |
| **Sleep disorders** |  |  |  |
| severity |  |  |  |
| 0 | 203 | 326 | 133 |
| 1 | 18 | 27 | 29 |
| 2 | 2 | 13 | 11 |
| 3 | 0 | 1 | 2 |
| **Depression** |  |  |  |
| Severity |  |  |  |
| 0 | 211 | 299 | 118 |
| 1 | 10 | 50 | 45 |
| 2 | 2 | 17 | 12 |
| 3 | 0 | 1 | 0 |
| **Appetite changes** |  |  |  |
| Severity |  |  |  |
| 0 | 222 | 326 | 147 |
| 1 | 1 | 29 | 21 |
| 2 | 0 | 11 | 7 |
| 3 | 0 | 1 | 0 |
| **Anxiety** |  |  |  |
| Severity |  |  |  |
| 0 | 215 | 304 | 119 |
| 1 | 5 | 49 | 41 |
| 2 | 3 | 13 | 14 |
| 3 | 0 | 1 | 1 |
| **Apathy** |  |  |  |
| Severity |  |  |  |
| 0 | 220 | 318 | 116 |
| 1 | 3 | 28 | 39 |
| 2 | 0 | 16 | 16 |
| 3 | 0 | 4 | 4 |

**Legend.** Symptom severity rating = 1, 2, 3

**ADNI Co-investigator**

Michael Weiner, MD (UC San Francisco, PI of ADNI), Paul Aisen, MD (UC San Diego, ADCS PI and Director of Coordinating Center Clinical Core), Michael Weiner, MD (UC San Francisco, Executive Committee), Paul Aisen, MD (UC San Diego, Executive Committee), Ronald Petersen, MD, PhD (Mayo Clinic, Rochester, Executive Committee), Clifford R. Jack, Jr., MD (Mayo Clinic, Rochester, Executive Committee), William Jagust, MD (UC Berkeley, Executive Committee), John Q. Trojanowki, MD, PhD (U Pennsylvania, Executive Committee), Arthur W. Toga, PhD (UCLA, Executive Committee), Laurel Beckett, PhD (UC Davis, Executive Committee), Robert C. Green, MD, MPH (Brigham and Women’s Hospital/Harvard Medical School, Executive Committee), Andrew J. Saykin, PsyD (Indiana University, Executive Committee), John Morris, MD (Washington University St. Louis, Executive Committee), Enchi Liu, PhD (Janssen Alzheimer Immunotherapy, ADNI 2 Private Partner Scientific Board (Chair), Robert C. Green, MD, MPH (Brigham and Women’s Hospital/Harvard Medical School, Data and Publication Committee (Chair)), Tom Montine, MD, PhD (University of Washington, Resource Allocation Review Committee), Ronald Petersen, MD, PhD (Mayo Clinic, Rochester, Clinical Core Leaders (Core PI)), Paul Aisen, MD (UC San Diego, Clinical Core Leaders), Anthony Gamst, PhD (UC San Diego, Clinical Informatics and Operations), Ronald G. Thomas, PhD (UC San Diego, Clinical Informatics and Operations), Michael Donohue, PhD (UC San Diego, Clinical Informatics and Operations), Sarah Walter, MSc (UC San Diego, Clinical Informatics and Operations), Devon Gessert (UC San Diego, Clinical Informatics and Operations), Tamie Sather (UC San Diego, Clinical Informatics and Operations), Laurel Beckett, PhD (UC Davis, Biostatistics Core Leaders and Key Personnel (Core PI)), Danielle Harvey, PhD (UC Davis, Biostatistics Core Leaders and Key Personnel), Anthony Gamst, PhD (UC San Diego, Biostatistics Core Leaders and Key Personnel), Michael Donohue, PhD (UC San Diego, Biostatistics Core Leaders and Key Personnel), John Kornak, PhD (UC Davis, Biostatistics Core Leaders and Key Personnel), Clifford R. Jack, Jr., MD (Mayo Clinic, Rochester, MRI Core Leaders and Key Personnel (Core PI)), Anders Dale, PhD (UC San Diego, MRI Core Leaders and Key Personnel), Matthew Bernstein, PhD (Mayo Clinic, Rochester, MRI Core Leaders and Key Personnel), Joel Felmlee, PhD (Mayo Clinic, Rochester, MRI Core Leaders and Key Personnel), Nick Fox, MD (University of London, MRI Core Leaders and Key Personnel), Paul Thompson, PhD (UCLA School of Medicine, MRI Core Leaders and Key Personnel), Norbert Schuff, PhD (UCSF MRI, MRI Core Leaders and Key Personnel), Gene Alexander, PhD (Banner Alzheimer’s Institute, MRI Core Leaders and Key Personnel), Charles DeCarli, MD (UC Davis, MRI Core Leaders and Key Personnel), William Jagust, MD (UC Berkeley, PET Core Leaders and Key Personnel (Core PI)), Dan Bandy, MS, CNMT (Banner Alzheimer’s Institute, PET Core Leaders and Key Personnel), Robert A. Koeppe, PhD (University of Michigan, PET Core Leaders and Key Personnel), Norm Foster, MD (University of Utah, PET Core Leaders and Key Personnel), Eric M. Reiman, MD (Banner Alzheimer’s Institute, PET Core Leaders and Key Personnel), Kewei Chen, PhD (Banner Alzheimer’s Institute, PET Core Leaders and Key Personnel), Chet Mathis, MD (University of Pittsburgh, PET Core Leaders and Key Personnel), John Morris, MD (Washington University St. Louis, Neuropathology Core Leaders), Nigel J. Cairns, PhD, (MRCPath Washington University St. Louis, Neuropathology Core Leaders), Lisa Taylor-Reinwald, BA, HTL (Washington University St. Louis, Neuropathology Core Leaders), J.Q. Trojanowki, MD, PhD (UPenn School of Medicine, Biomarkers Core Leaders and Key Personnel (Core PI)), Les Shaw, PhD (UPenn School of Medicine, Biomarkers Core Leaders and Key Personnel), Virginia M.Y. Lee, PhD, MBA (UPenn School of Medicine, Biomarkers Core Leaders and Key Personnel), Magdalena Korecka, PhD (UPenn School of Medicine, Biomarkers Core Leaders and Key Personnel), Arthur W. Toga, PhD (UCLA, Informatics Core Leaders and Key Personnel (Core PI)), Karen Crawford (UCLA, Informatics Core Leaders and Key Personnel), Scott Neu, PhD (UCLA, Informatics Core Leaders and Key Personnel), Andrew J. Saykin, PsyD (Indiana University, Genetics Core Leaders and Key Personnel), Tatiana M. Foroud, PhD (Indiana University, Genetics Core Leaders and Key Personnel), Steven Potkin, MD (UC Irvine, Genetics Core Leaders and Key Personnel), Li Shen, PhD (Indiana University, Genetics Core Leaders and Key Personnel), Zaven Kachaturian, PhD (Khachaturian, Radebaugh & Associates (KRA), Inc, Early Project Development), Richard Frank, MD, PhD (General Electric, Early Project Development), Peter J. Snyder, PhD (University of Connecticut, Early Project Development), Susan Molchan, PhD (National Institute on Aging/National Institutes of Health, Early Project Development), Jeffrey Kaye, MD (Oregon Health and Science University, Site Investigator), Joseph Quinn, MD (Oregon Health and Science University, Site Investigator), Betty Lind, BS (Oregon Health and Science University, Site Investigator), Sara Dolen, BS (Oregon Health and Science University, Past Site Investigator), Lon S. Schneider, MD (University of Southern California, Site Investigator), Sonia Pawluczyk, MD (University of Southern California, Site Investigator), Bryan M. Spann, DO, PhD (University of Southern California, Site Investigator), James Brewer, MD, PhD (University of California--San Diego, Site Investigator), Helen Vanderswag, RN (University of California--San Diego, Site Investigator), Judith L. Heidebrink, MD, MS, (University of Michigan, Site Investigator), Joanne L. Lord, LPN, BA, CCRC (University of Michigan, Site Investigator), Ronald Petersen, MD, PhD (Mayo Clinic, Rochester, Site Investigator), Kris Johnson, RN (Mayo Clinic, Rochester, Site Investigator), Rachelle S. Doody, MD, PhD (Baylor College of Medicine, Site Investigator), Javier Villanueva-Meyer, MD (Baylor College of Medicine, Site Investigator), Munir Chowdhury, MBBS, MS (Baylor College of Medicine, Site Investigator), Yaakov Stern, PhD (Columbia University Medical Center, Site Investigator), Lawrence S. Honig, MD, PhD (Columbia University Medical Center, Site Investigator), Karen L. Bell, MD, (Columbia University Medical Center, Site Investigator), John C. Morris, MD (Washington University, St. Louis, Site Investigator), Beau Ances, MD (Washington University, St. Louis, Site Investigator), Maria Carroll, RN, MSN (Washington University, St. Louis, Site Investigator), Sue Leon, RN, MSN (Washington University, St. Louis, Site Investigator), Mark A. Mintun, MD (Washington University, St. Louis, Past Site Investigator), Stacy Schneider, APRN, BC, GNP (Washington University, St. Louis, Past Site Investigator), Daniel Marson, JD, PhD (University of Alabama – Birmingham, Site Investigator), Randall Griffith, PhD, ABPP (University of Alabama – Birmingham, Site Investigator), David Clark, MD (University of Alabama – Birmingham, Site Investigator), Hillel Grossman, MD (Mount Sinai School of Medicine, Site Investigator), Effie Mitsis, PhD (Mount Sinai School of Medicine, Site Investigator), Aliza Romirowsky, BA (Mount Sinai School of Medicine, Site Investigator), Leyla deToledo-Morrell, PhD (Rush University Medical Center, Site Investigator), Raj C. Shah, MD (Rush University Medical Center, Site Investigator) (Wein Center, Site Investigator), Ranjan Duara, MD (Wein Center, Site Investigator), Daniel Varon, MD (Wein Center, Site Investigator), Peggy Roberts, CNA (Wein Center, Site Investigator), Marilyn Albert, PhD (Johns Hopkins University, Site Investigator), Chiadi Onyike, MD, MHS (Johns Hopkins University, Site Investigator), Stephanie Kielb MD (Johns Hopkins University, Site Investigator), Henry Rusinek, PhD (New York University, Site Investigator), Mony J de Leon, EdD (New York University, Site Investigator), Lidia Glodzik, MD, PhD (New York University, Site Investigator), Susan De Santi, PhD (New York University, Past Site Investigator), P. Murali Doraiswamy, MD (Duke University Medical Center, Site Investigator), Jeffrey R. Petrella, MD (Duke University Medical Center, Site Investigator), R. Edward Coleman, MD (Duke University Medical Center, Site Investigator), Steven E. Arnold, MD (University of Pennsylvania, Site Investigator), Jason H. Karlawish, MD, (University of Pennsylvania, Site Investigator), David Wolk, MD (University of Pennsylvania, Site Investigator), Charles D. Smith, MD (University of Kentucky, Site Investigator), Greg Jicha, MD (University of Kentucky, Site Investigator), Peter Hardy, PhD (University of Kentucky, Site Investigator), Oscar L. Lopez, MD (University of Pittsburgh, Site Investigator), MaryAnn Oakley, MA (University of Pittsburgh, Site Investigator), Donna M. Simpson, CRNP, MPH (University of Pittsburgh, Site Investigator), Anton P. Porsteinsson, MD (University of Rochester Medical Center, Site Investigator), Bonnie S. Goldstein, MS, NP (University of Rochester Medical Center, Site Investigator), Kim Martin, RN (University of Rochester Medical Center, Site Investigator), Kelly M. Makino, BS (University of Rochester Medical Center, Past Site Investigator), M. Saleem Ismail, MD (University of Rochester Medical Center, Past Site Investigator), Connie Brand, RN (University of Rochester Medical Center, Past Site Investigator), Ruth A. Mulnard, DNSc, RN, FAAN (University of California, Irvine, Site Investigator), Gaby Thai, MD (University of California, Irvine, Site Investigator), Catherine Mc-Adams-Ortiz, MSN, RN, A/GNP (University of California, Irvine, Site Investigator), Kyle Womack, MD (University of Texas Southwestern Medical School, Site Investigator), Dana Mathews, MD, PhD (University of Texas Southwestern Medical School, Site Investigator), Mary Quiceno, MD (University of Texas Southwestern Medical School, Site Investigator), Ramon Diaz-Arrastia, MD, PhD (University of Texas Southwestern Medical School, Past Site Investigator), Richard King, MD (University of Texas Southwestern Medical School, Past Site Investigator), Myron Weiner, MD (University of Texas Southwestern Medical School, Past Site Investigator), Kristen Martin-Cook, MA (University of Texas Southwestern Medical School, Past Site Investigator), Michael DeVous, PhD (University of Texas Southwestern Medical School, Past Site Investigator), Allan I. Levey, MD, PhD (Emory University, Site Investigator), James J. Lah, MD, PhD (Emory University, Site Investigator), Janet S. Cellar, DNP, PMHCNS-BC (Emory University, Site Investigator), Jeffrey M. Burns, MD (University of Kansas, Medical Center, Site Investigator), Heather S. Anderson, MD (University of Kansas, Medical Center, Site Investigator), Russell H. Swerdlow, MD (University of Kansas, Medical Center, Site Investigator), Liana Apostolova, MD (University of California, Los Angeles, Site Investigator), Po H. Lu, PsyD (University of California, Los Angeles, Site Investigator), George Bartzokis, MD (University of California, Los Angeles, Past Site Investigator), Daniel H.S. Silverman, MD, PhD (University of California, Los Angeles, Past Site Investigator), Neill R Graff-Radford, MBBCH, FRCP (London) (Mayo Clinic, Jacksonville, Site Investigator), Francine Parfitt, MSH, CCRC (Mayo Clinic, Jacksonville, Site Investigator), Heather Johnson, MLS, CCRP (Mayo Clinic, Jacksonville, Site Investigator), Martin R. Farlow, MD (Indiana University, Site Investigator), Ann Marie Hake, MD, Brandy R. Matthews, MD (Indiana University, Site Investigator), Scott Herring, RN (Indiana University, Past Site Investigator), Christopher H. van Dyck, MD (Yale University School of Medicine, Site Investigator), Richard E. Carson, PhD (Yale University School of Medicine, Site Investigator), Martha G. MacAvoy, PhD (Yale University School of Medicine, Site Investigator), Howard Chertkow, MD (McGill Univ., Montreal-Jewish General Hospital, Site Investigator), Howard Bergman, MD (McGill Univ., Montreal-Jewish General Hospital, Site Investigator), Chris Hosein, MEd (McGill Univ., Montreal-Jewish General Hospital, Site Investigator), Sandra Black, MD, FRCPC (Sunnybrook Health Sciences, Ontario, Site Investigator), Bojana Stefanovic, PhD (Sunnybrook Health Sciences, Ontario, Site Investigator), Curtis Caldwell, PhD (Sunnybrook Health Sciences, Ontario, Site Investigator), Ging-Yuek Robin Hsiung, MD, MHSc, FRCPC (U.B.C. Clinic for AD & Related Disorders, Site Investigator), Howard Feldman, MD, FRCPC (U.B.C. Clinic for AD & Related Disorders, Site Investigator), Benita Mudge, BS (U.B.C. Clinic for AD & Related Disorders, Site Investigator), Michele Assaly, MA (U.B.C. Clinic for AD & Related Disorders, Past Site Investigator), Andrew Kertesz, MD (Cognitive Neurology - St. Joseph's, Ontario, Site Investigator), John Rogers, MD (Cognitive Neurology - St. Joseph's, Ontario, Site Investigator), Dick Trost, PhD (Cognitive Neurology - St. Joseph's, Ontario, Site Investigator), Charles Bernick, MD (Cleveland Clinic Lou Ruvo Center for Brain Health, Site Investigator), Donna Munic, PhD (Cleveland Clinic Lou Ruvo Center for Brain Health, Site Investigator), Diana Kerwin, MD (Northwestern University, Site Investigator), Marek-Marsel Mesulam, MD (Northwestern University, Site Investigator), Kristina Lipowski, BA (Northwestern University, Site Investigator), Chuang-Kuo Wu, MD, PhD (Northwestern University, Past Site Investigator), Nancy Johnson, PhD (Northwestern University, Past Site Investigator), Carl Sadowsky, MD (Premiere Research Inst (Palm Beach Neurology), Site Investigator), Walter Martinez, MD (Premiere Research Inst (Palm Beach Neurology), Site Investigator), Teresa Villena, MD (Premiere Research Inst (Palm Beach Neurology), Site Investigator), Raymond Scott Turner, MD, PhD (Georgetown University Medical Center, Site Investigator), Kathleen Johnson, NP (Georgetown University Medical Center, Site Investigator), Brigid Reynolds, NP (Georgetown University Medical Center, Site Investigator), Reisa A. Sperling, MD (Brigham and Women's Hospital, Site Investigator), Keith A. Johnson, MD (Brigham and Women's Hospital, Site Investigator), Gad Marshall, MD (Brigham and Women's Hospital, Past Site Investigator), Meghan Frey (Brigham and Women's Hospital, Past Site Investigator), Jerome Yesavage, MD (Stanford University, Site Investigator), Joy L. Taylor, PhD (Stanford University, Site Investigator), Barton Lane, MD (Stanford University, Site Investigator), Allyson Rosen, PhD (Stanford University, Past Site Investigator), Jared Tinklenberg, MD (Stanford University, Past Site Investigator), Marwan Sabbagh, MD, FAAN, CCRI (Banner Sun Health Research Institute, Site Investigator), Christine Belden, PsyD (Banner Sun Health Research Institute, Site Investigator), Sandra Jacobson, MD (Banner Sun Health Research Institute, Site Investigator), Neil Kowall, MD (Boston University, Site Investigator), Ronald Killiany, PhD (Boston University, Site Investigator), Andrew E. Budson, MD (Boston University, Site Investigator), Alexander Norbash, MD (Boston University, Past Site Investigator), Patricia Lynn Johnson, BA (Boston University, Past Site Investigator), Thomas O. Obisesan, MD, MPH (Howard University, Site Investigator), Saba Wolday, MSc (Howard University, Site Investigator), Salome K. Bwayo, PharmD (Howard University, Past Site Investigator), Alan Lerner, MD (Case Western Reserve University, Site Investigator), Leon Hudson, MPH (Case Western Reserve University, Site Investigator), Paula Ogrocki, PhD (Case Western Reserve University, Site Investigator), Evan Fletcher, PhD (University of California, Davis – Sacramento, Site Investigator), Owen Carmichael, PhD (University of California, Davis – Sacramento, Site Investigator), John Olichney, MD (University of California, Davis – Sacramento, Site Investigator), Charles DeCarli, MD (University of California, Davis – Sacramento, Past Site Investigator), Smita Kittur, MD (Neurological Care of CNY, Site Investigator), Michael Borrie, MB ChB (Parkwood Hospital, Site Investigator), T-Y Lee, PhD (Parkwood Hospital, Site Investigator), Dr Rob Bartha, PhD (Parkwood Hospital, Site Investigator), Sterling Johnson, PhD (University of Wisconsin, Site Investigator), Sanjay Asthana, MD (University of Wisconsin, Site Investigator), Cynthia M. Carlsson, MD (University of Wisconsin, Site Investigator), Steven G. Potkin, MD (University of California, Irvine – BIC, Site Investigator), Adrian Preda, MD (University of California, Irvine – BIC, Site Investigator), Dana Nguyen, PhD (University of California, Irvine – BIC, Site Investigator), Pierre Tariot, MD (Banner Alzheimer's Institute, Site Investigator), Adam Fleisher, MD (Banner Alzheimer's Institute, Site Investigator), Stephanie Reeder, BA (Banner Alzheimer's Institute, Site Investigator), Vernice Bates, MD (Dent Neurologic Institute, Site Investigator), Horacio Capote, MD (Dent Neurologic Institute, Site Investigator), Michelle Rainka, PharmD, CCRP (Dent Neurologic Institute, Site Investigator), Douglas W. Scharre, MD (Ohio State University, Site Investigator), Maria Kataki, MD, PhD (Ohio State University, Site Investigator), Earl A. Zimmerman, MD (Albany Medical College, Site Investigator), Dzintra Celmins, MD (Albany Medical College, Site Investigator), Alice D. Brown, FNP (Albany Medical College, Past Site Investigator), Godfrey D. Pearlson, MD (Hartford Hosp, Olin Neuropsychiatry Research Center, Site Investigator), Karen Blank, MD (Hartford Hosp, Olin Neuropsychiatry Research Center, Site Investigator), Karen Anderson, RN (Hartford Hosp, Olin Neuropsychiatry Research Center, Site Investigator), Andrew J. Saykin, PsyD (Dartmouth-Hitchcock Medical Center, Site Investigator), Robert B. Santulli, MD (Dartmouth-Hitchcock Medical Center, Site Investigator), Eben S. Schwartz, PhD (Dartmouth- Hitchcock Medical Center, Site Investigator), Kaycee M. Sink, MD, MAS (Wake Forest University Health Sciences, Site Investigator), Jeff D. Williamson, MD, MHS (Wake Forest University Health Sciences, Site Investigator), Pradeep Garg, PhD (Wake Forest University Health Sciences, Site Investigator), Franklin Watkins, MD (Wake Forest University Health Sciences, Past Site Investigator), Brian R. Ott, MD (Rhode Island Hospital, Site Investigator), Henry Querfurth, MD (Rhode Island Hospital, Site Investigator), Geoffrey Tremont, PhD (Rhode Island Hospital, Site Investigator), Stephen Salloway, MD, MS (Butler Hospital, Site Investigator), Paul Malloy, PhD (Butler Hospital, Site Investigator), Stephen Correia, PhD (Butler Hospital, Site Investigator), Howard J. Rosen, MD (UC San Francisco, Site Investigator), Bruce L. Miller, MD (UC San Francisco, Site Investigator), Jacobo Mintzer, MD, MBA (Medical University South Carolina, Site Investigator), Crystal Flynn Longmire, PhD (Medical University South Carolina, Site Investigator), Kenneth Spicer, MD, PhD (Medical University South Carolina, Site Investigator), Elizabether Finger, MD (St. Joseph’s Health Care, Site Investigator), Irina Rachinsky, MD (St. Joseph’s Health Care, Site Investigator), John Rogers, MD (St. Joseph’s Health Care, Site Investigator), Andrew Kertesz, MD (St. Joseph’s Health Care, Past Site Investigator), Dick Drost, MD (St. Joseph’s Health Care, Past Site Investigator), Nunzio Pomara, MD (Nathan Kline Institute, Site Investigator), Raymundo Hernando, MD (Nathan Kline Institute, Site Investigator), Antero Sarrael, MD (Nathan Kline Institute, Site Investigator), Susan K. Schultz, MD (University of Iowa, Site Investigator), Laura L. Boles Ponto, PhD (University of Iowa, Site Investigator), Hyungsub Shim, MD (University of Iowa, Site Investigator), Karen Elizabeth Smith, RN (University of Iowa, Site Investigator), Norman Relkin, MD, PhD (Cornell University), Gloria Chaing, MD (Cornell University), Lisa Raudin, PhD (Cornell University), Amanda Smith, MD (University of South Florida), Kristin Fargher, MD (University of South Florida), Balebail Ashok Raj, MD (University of South Florida).
